# Clinicopathologic correlates of pembrolizumab efficacy in patients with advanced NSCLC and a PD-L1 expression of ≥ 50%

**DOI:** 10.1101/2020.04.09.20047464

**Authors:** Alessio Cortellini, Marcello Tiseo, Giuseppe L Banna, Federico Cappuzzo, Joachim GJV Aerts, Fausto Barbieri, Raffaele Giusti, Emilio Bria, Diego Cortinovis, Francesco Grossi, Maria R Migliorino, Domenico Galetta, Francesco Passiglia, Daniele Santini, Rossana Berardi, Alessandro Morabito, Carlo Genova, Francesca Mazzoni, Vincenzo Di Noia, Diego Signorelli, Alessandro Tuzi, Alain Gelibter, Paolo Marchetti, Marianna Macerelli, Francesca Rastelli, Rita Chiari, Danilo Rocco, Stefania Gori, Michele De Tursi, Giovanni Mansueto, Federica Zoratto, Matteo Santoni, Marianna Tudini, Erika Rijavec, Marco Filetti, Annamaria Catino, Pamela Pizzutilo, Luca Sala, Fabrizio Citarella, Russano Marco, Mariangela Torniai, Luca Cantini, Giada Targato, Vincenzo Sforza, Olga Nigro, Miriam G Ferrara, Ettore D’Argento, Sebastiano Buti, Paola Bordi, Lorenzo Antonuzzo, Simona Scodes, Lorenza Landi, Giorgia Guaitoli, Cinzia Baldessari, Luigi Della Gravara, Maria Giovanna Dal Bello, Robert A. Belderbos, Paolo Bironzo, Simona Carnio, Serena Ricciardi, Alessio Grieco, Alessandro De Toma, Claudia Proto, Alex Friedlaender, Ornella Cantale, Biagio Ricciuti, Alfredo Addeo, Giulio Metro, Corrado Ficorella, Giampiero Porzio

## Abstract

**Background:** Single agent pembrolizumab represents the standard first line option for metastatic non-small-cell-lung-cancer (NSCLC) patients with a PD-L1 (programmed death-ligand 1) expression of ≥ 50%.

**Methods:** We conducted a multicenter study aimed at evaluating the clinicopathologic correlates of pembrolizumab efficacy in patients with treatment-naïve NSCLC and a PD-L1 TPS ≥ 50%.

**Results:** 1026 consecutive patients were included. ECOG-PS ≥ 2 (p < 0.0001) and bone metastases (p = 0.0003) were confirmed to be independent predictors of a worse ORR. Former smokers (p = 0.0002), but not current smokers (p = 0.0532) were confirmed to have a significantly prolonged PFS compared to never smokers at multivariate analysis. ECOG-PS (p < 0.0001), bone metastases (p < 0.0001) and liver metastases (p < 0.0001) were also confirmed to be independent predictors of a worse PFS. Previous palliative RT was significantly related to a shortened OS (p = 0.0104), while previous non-palliative RT was significantly related to a prolonged OS (p = 0.0033). Former smokers (p = 0.0131), but not current smokers (p = 0.3433) were confirmed to have a significantly prolonged OS compared to never smokers. ECOG-PS (p < 0.0001), bone metastases (p < 0.0001) and liver metastases (p < 0.0001) were also confirmed to be independent predictors of a shortened OS. A PD-L1 expression of ≥ 90%, as assessed by recursive partitioning, was associated with significantly higher ORR (p = 0.0204), and longer and OS (p = 0.0346) at multivariable analysis.

**Conclusions:** pembrolizumab was effective in a large cohort of NSCLC patients treated outside of clinical trials. We confirmed that the absence of tobacco exposure, and the presence of bone and liver metastasis are associated with worse clinical outcomes to pembrolizumab. Increasing levels of PD-L1 expression may help identifying a subset of patients who derive a greater benefit from pembrolizumab monotherapy.

## Introduction

Based on the results of the Keynote-024 trial, single agent pembrolizumab has become the standard of care for the first line treatment of metastatic non-small-cell-lung-cancer (NSCLC) patients with a tumor proportion score (TPS) of PD-L1 (programmed death-ligand 1) ≥ 50%, lacking *EGFR* mutation and *ALK* rearrangement [1-3]. However, clinical trials data often do not apply to real life populations. Recent real-world experiences with pembrolizumab monotherapy showed inferior progression free survival (PFS) and overall survival (OS), compared to the Keynote-024 experimental arm [4-6], as patients with poor performance status and genetic drivers are usually excluded form clinical trials. For instance, a retrospective multicenter study of pembrolizumab monotherapy in NSCLC patients, with PD-L1 TPS ≥ 50% and Eastern Cooperative Performance Status (ECOG-PS) ≥ 2, has revealed poor clinical outcomes to treatment, particularly in those patients whose poor PS was related to a high disease burden [7].

More recently, the Keynote-189 and Keynote-407 trials have shown that the addition of pembrolizumab to a platinum-based chemotherapy, improved clinical outcomes to placebo, in both the adenocarcinoma and squamous NSCLC, regardless of PD-L1 expression [8-9]. However, the subgroups analyses according to PD-L1 expression levels, confirmed that the efficacy was higher among patients with PD-L1 TPS ≥ 50% [8-9]. However, the efficacy of pembrolizumab monotherapy in patients treated outside of clinical trials is still in need of further investigation. To address this need, we conducted this multicenter study, aimed at evaluating clinicopathologic correlates of pembrolizumab monotherapy in NSCLC patients with a PD-L1 expression of ≥ 50%, in a large real-life cohort.

## Materials and Methods

### Study Design

This multicenter retrospective study evaluated metastatic NSCLC patients with PD-L1 TPS of ≥ 50%, consecutively treated with first line pembrolizumab monotherapy, from January 2017 to October 2019, at 34 institutions (Supplementary file 1). The sample size was estimated according to the expected enrollment of the participating centers.

The primary aim of this analysis was to describe clinical outcome of metastatic NSCLC patients receiving pembrolizumab monotherapy in clinical practice. The measured clinical outcomes were objective response rate (ORR), median progression-free survival (PFS) and median overall survival (OS). Secondly, to evaluate whether some baseline clinical factors affected clinical outcomes, univariate and multivariate analyses of ORR, PFS and OS were performed (using a stepwise selection of covariates, with an entry significance level of 0.05).

Patients were assessed with radiological imaging according to the local clinical practice; RECIST (v. 1.1) criteria were used [10], but treatment beyond disease progression was allowed when clinically indicated. ORR was defined as the portion of patients experiencing an objective response (complete or partial response) as best response to immunotherapy. PFS was defined as the time from treatment’s start to disease progression or death whichever occurred first; OS as the time from the beginning of treatment to death. The analyzed clinical factors in the univariate/multivariate analyses were:

- PD-L1 expression (< *vs*. ≥ the computed optimal cut off for);
- Smoking status (never smokers *vs*. former smokers [≥ 1 year]/current smokers) [11];
- Age (< 70 *vs*. ≥ 70 years old) [12];
- Sex (male *vs* female);
- ECOG-PS (0-1 *vs* ≥ 2);
- Histology (Squamous *vs*.Non-squamous [including mixed hisologies]);
- Central Nervous System (CNS) metastases (yes *vs* no);
- Bone metastases (yes *vs* no);
- Liver metastases (yes *vs* no);
- Corticosteroids administration (dose equivalent or higher to 10 mg prednisone per day) within the 30 days before treatment commencement (named baseline steroids) (yes *vs* no);
- Radiation therapy (RT) within the previous 6 months the immunotherapy commencement (no RT *vs*. non-palliative RT [e.g. single-fraction stereotactic radiosurgery, stereotactic RT to a metastatic site]/palliative RT [e.g. whole brain radiation therapy, and any other treatment administered for symptoms palliation and/or without a curative intent]) [13];

In order to properly weighing the role of baseline clinical factors, and to find appropriate covariates to be used in the multivariate models, the correlations between baseline steroids, previous RT and disease burden (CNS metastases, bone metastases and liver metastases) were evaluated with the χ2 test and χ2 test for trend [14]. In case of a significant relationship, they were not used in the multivariate analyses [15]. The χ2 test and χ2 test for trend were also used to compare ORR among subgroups [14]; logistic regression was used for the multivariate analysis of ORR, and adjusted odds ratios (ORs) with 95% confidence intervals (95%CI) were computed [16]. Median PFS and median OS were evaluated using the Kaplan-Meier method [17]. Median period of follow-up was computed according to the reverse Kaplan-Meier method [18]. Cox proportional hazards regression was used to evaluate predictor variables and estimate the hazard ratios (HRs) for PFS and OS [19]. Data cut off period was February 2020. All statistical analyses were performed using MedCalc Statistical Software version 18.11.3 (MedCalc Software bvba, Ostend, Belgium; https://www.medcalc.org; 2019). Recursive partitioning was performed using the R package rpart (R version 3.6.2).

### PD-L1 TPS evaluation

PD-L1 expression was reported as a percentage of tumor cells with positive membranous staining, using a variety of immunohistochemical antibodies and platforms according to local institutional clinical practice (including the 22C3, SP263, E1L3N, and 28-8 antibodies). Being the TPS evaluation validated only with the 22C3 [20], we referred to “PD-L1 expression” in our study.

To determine whether among patients with a PD-L1 expression ranging from 50% to 100%, increasing levels of PD-L1 were predictive of pembrolizumab efficacy, a ROC curve analyses of ORR was performed [21]. As complementary analysis, to identify an optimal grouping according to PD-L1 expression, with respect to ORR, PFS and OS, a recursive partitioning algorithm was used, using the Rpart function in R, as previously done [22-23]. As in clinical practice the PD-L1 TPS of some patients was reported as “≥ 50%”, and not as discrete value, we included in this analysis only the patients with data availability regarding the absolute estimated value of PD-L1 TPS.

Even considering the concordance between metachronous and synchronous formalin-fixed, paraffin-embedded (FFPE) tissue for PD-L1 estimation, and the growing data suggesting the reliability of non-FFPE samples such as liquid-based cytology [23], a one-way analysis of variance of PD-L1 expression according to the tissue specimen type was performed [24]. Tissue specimens were categorized in surgical samples (both metachronous and synchronous), tissue biopsies and cytological specimens. A one-way analysis of variance of PD-L1 expression according to the smoking status was also performed.

## Results

### Patients characteristics

One thousand and twenty-six consecutive metastatic NSCLC patients, with PD-L1 TPS ≥ 50%, were included. Patient characteristics and RT details are summarized in Table 1. As reported in Table 2, baseline steroids and previous RT were significantly related with CNS metastases (p < 0.0001 and p < 0.0001, respectively) and bone metastases (p < 0.0001 and p = 0.0389, respectively). Baseline steroids were also significantly related to liver metastases (p = 0.0225) No significant associations were found between previous RT and liver metastases. Among the 628 patients (61.2%) who discontinued first line pembrolizumab at the data cut-off, only 200 patients (31.8%) underwent a second line disease-oriented treatment, while among the 599 patients (58.4%) who experienced disease progression, 428 patients (71.5%) were deceased. Among the 428 deceased patients, 332 (77.6%) did not received a second line disease-oriented treatment.

**Table 1:** Patients characteristics. *Available for 731 patients (67.3%). # Available for 128 out of 181 (70.7%) palliative radiation treatments. ¥ Available for 35 out of 47 (74.5%) non-palliative radiation treatments. NA: not available

|  | N° (%) |
| --- | --- |
|  | **1026** |
| **AGE, (years)** |  |
| Median | 70.2 |
| Range | 28 – 92 |
| Elderly ($\geq 70$) | 529 (51.6) |
| **Smoking status** |  |
| Never smokers | 106 (10.3) |
| Former smokers | 572 (55.8) |
| Current smokers | 348 (33.9) |
| **SEX** |  |
| Male | 673 (65.6) |
| Female | 353 (34.4) |
| **ECOG PS** |  |
| 0 - 1 | 847 (82.6) |
| $\geq 2$ | 179 (17.4) |
| **Histology** |  |
| Squamous | 248 (24.2) |
| Non-squamous | 778 (75.8) |
| **Tissue specimen** |  |
| Surgical sample | 84 (8.2) |
| Tissue biopsy | 769 (75.0) |
| Cytological specimen | 109 (10.6) |
| NA | 64 (6.2) |
| **PD-L1 antibody clone** |  |
| 22C3 | 620 (60.4) |
| SP263 | 329 (32.1) |
| E1L3N | 9 (0.9) |
| 28-8 | 18 (1.7) |
| NA | 51 (4.9) |
| **PD-L1 expression (%)\*** |  |
| Mean | 72.2 |
| Median | 70 |
| Range | (50 – 100) |
| **CNS metastases** |  |
| Yes | 181 (17.6) |
| No | 845 (82.4) |
| **Bone metastases** |  |
| Yes | 326 (31.8) |
| No | 700 (68.2) |
| **Liver metastases** |  |
| Yes | 158 (15.4) |
| No | 868 (84.6) |
| **Baseline steroids** |  |
| Yes | 251 (24.5) |
| No | 757 (75.5) |
| **Previous RT** |  |
| No RT | 814 (79.3) |
| Non-palliative | 44 (4.3) |
| Palliative | 168 (16.4) |
| **EGFR mutated patients** | **10 Patients (0.9)** |
| Exon 20 mutations | 4 (40) |
| Exon 19 deletions | 4 (40) |
| Exon 21 mutation | 1 (10) |
| NA | 1 (10) |
| **Palliative radiation treatments** | **181 treatments** |
| Bone | 107 (59.1) |
| CNS | 53 (29.3) |
| Lymph nodes | 8 (4.4) |
| Lung/chest wall | 9 (5.0) |
| Others | 4 (2.2) |
| Median dose Gray (range)# | 20 (8 – 40) |
| **Non-palliative radiation treatments** | **47 treatments** |
| CNS | 36 (76.6) |
| Non-CNS | 11 (23.4) |
| Median dose Gray (range)¥ | 24.5 (15 – 60) |

**Table 2:**
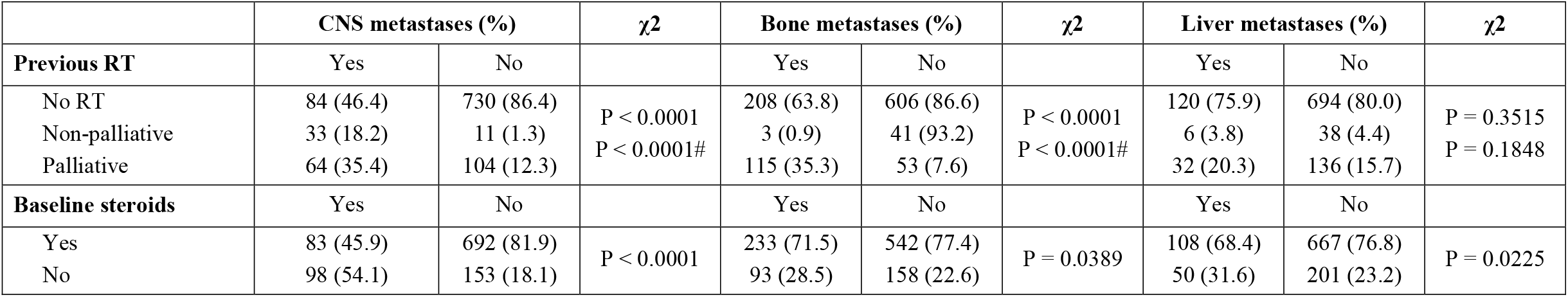
Correlation analyses between previous RT categories, baseline steroids and sites of metastases. # χ2 test for trend.

| | CNS metastases (%) | | $\chi^2$ | Bone metastases (%) | | $\chi^2$ | Liver metastases (%) | | $\chi^2$ |
| --- | --- | --- | --- | --- | --- | --- | --- | --- | --- |
| Previous RT | Yes | No |  | Yes | No |  | Yes | No |  |
| No RT | 84 (46.4) | 730 (86.4) | P < 0.0001<br>P < 0.0001# | 208 (63.8) | 606 (86.6) | P < 0.0001<br>P < 0.0001# | 120 (75.9) | 694 (80.0) | P = 0.3515<br>P = 0.1848 |
| Non-palliative | 33 (18.2) | 11 (1.3) |  | 3 (0.9) | 41 (93.2) |  | 6 (3.8) | 38 (4.4) |  |
| Palliative | 64 (35.4) | 104 (12.3) |  | 115 (35.3) | 53 (7.6) |  | 32 (20.3) | 136 (15.7) |  |
| Baseline steroids | Yes | No |  | Yes | No |  | Yes | No |  |
| Yes | 83 (45.9) | 692 (81.9) | P < 0.0001 | 233 (71.5) | 542 (77.4) | P = 0.0389 | 108 (68.4) | 667 (76.8) | P = 0.0225 |
| No | 98 (54.1) | 153 (18.1) |  | 93 (28.5) | 158 (22.6) |  | 50 (31.6) | 201 (23.2) |  |

### PD-L1 analysis

The ROC curve analysis for PD-L1 TPS of ORR revealed a weak predictive performance within the range 50%-100% (AUC = 0.55 [95%CI: 0.51-0.59], p = 0.0303) (Figure 1A). The absolute value of PD-L1 TPS was available for 731 patients (71.2%) and the median TPS was 70%. The mean PD-L1 TPS for cytological specimens, surgical samples and tissue biopsies were 69% (standard deviation [sd]: 14), 68% (sd: 13) and 73% (sd: 13) respectively; the analysis of variance showed that the tissue specimen type significantly affected the PD-L1 TPS evaluation [F (2,668) = 3.19, p = 0.042]. The mean PD-L1 TPS of current smokers, former smokers and never smokers were 73% (sd: 14), 72% (sd: 14) and 70% (sd: 11), respectively; smoking status significantly affected the PD-L1 TPS estimation [F (2,728) = 0.92, p = 0.008]. Figure 1B and Figure 1C reported the respective multiple comparison graph. The recursive partitioning algorithm however identified a primary split at a PD-L1 expression level of 91.9% (p = 0.019) and 92% (p = 0.022) for ORR and OS, respectively. No significant splits were found regarding PFS. We therefore used the optimal grouping cut-off of 90% for clinical outcome analysis.

**Figure 1:**
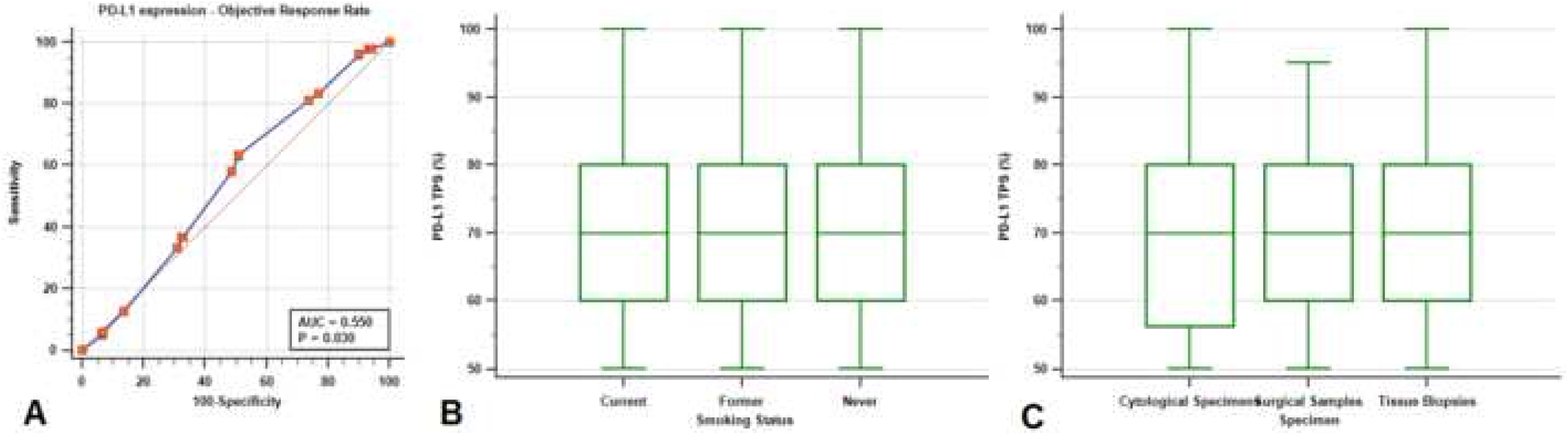
Multiple comparison graphs of PD-L1 TPS according to the smoking status (A) and to the tissue specimen (B).

### Clinical outcomes analysis

In the entire study population, the ORR was 44.5% (95%CI: 40.2-49.1% [899 evaluable patients for ORR]). Table 3 summarizes the univariate and multivariate analysis of ORR. The use of baseline steroids was significantly related to inferior ORR at the univariate analysis (29.9% vs 48.7%, p < 0.0001). A PD-L1 TPS of <90%, ECOG-PS ≥ 2 and baseline bone metastases were confirmed to be independent predictors of a worse ORR. At the data cut off the median follow-up was 14.6 months (95%CI: 13.5-15.6). The median PFS and median OS of the study population were 7.9 months (95%CI: 6.9-9.5) and 17.2 months (95%CI: 15.3-22.3), respectively. The median PFS of patients with PD-L1 expression < 90% was 6.9 months (95%CI: 5.8-8.4), which was significantly shorter compared the PFS of 12.0 (95%CI: 6.3-19.4) months of patients with a PD-L1 expression of ≥ 90% (unadjusted HR = 1.29 [95%CI: 1.01–1.66], p=0.0487) (Figure 2A). PD-L1 expression was not confirmed an independent predictor for PFS at the multivariate analysis (Table 4). The median OS was also significantly shorter among patients with PD-L1 expression of < 90% as compared to those with a PD-L1 expression of ≥ 90% (14.7 months [95%CI: 11.1-17.3] months versus not reached, HR = 1.51 [95%CI: 1.10–2.07], p=0.0093) (Figure 2B). A PD-L1 TPS of <90% was confirmed an independent predictor for shorter OS at the multivariate analysis (Table 5).

**Table 3:** Univariate and multivariate analysis of ORR. *Available for 650 patients, not used in the multivariate analysis of the overall study population; ECOG-PS (≥ 2 *vs*. 0-1), bone metastases (yes *vs*. no) and liver metastases (yes *vs*. no) were used as adjusting factors for PD-L1 analysis. # χ2 test for trend.

|  | UNIVARIATE ANALYSIS |  |  | MULTIVARIATE |  |  |  |
| --- | --- | --- | --- | --- | --- | --- | --- |
| Variable (comparator) | Response/ Ratio | ORR (95% CI) | *p* - value | Coeff. | St. Err. | *p* - value | Adjusted OR (95% CI) |
| Overall | 400/899 | 44.5 (40.2–49.1) | - | - | - | - | - |
| PD-L1 expression* |  |  |  |  |  |  |  |
| < 90% | 224/524 | 42.7 (37.3–48.7) | 0.0347 | -0.0137 | 0.0059 | 0.0204 | 0.98 (0.97-0.99) |
| ≥ 90% | 67/126 | 53.2 (41.2–67.5) |  |  |  |  |  |
| Smoking status |  |  |  |  |  |  |  |
| (Never smoker) | 32/92 | 34.8 (23.8–49.1) | 0.0791 | - | - | - | - |
| Former smoker | 239/508 | 47.0 (42.2–53.4) |  |  |  |  |  |
| Current smoker | 129/299 | 43.1 (36.0–51.2) | 0.5948 |  |  |  |  |
| Sex |  |  |  |  |  |  |  |
| Male | 263/592 | 44.4 (39.2-50.1) | 0.9545 | - | - | - | - |
| Female | 137/307 | 44.6 (37.4-52.7) |  |  |  |  |  |
| Age |  |  |  |  |  |  |  |
| Elderly | 209/461 | 45.3 (39.4–51.9) | 0.6023 | - | - | - | - |
| Non-elderly | 191/438 | 43.6 (37.6-50.2) |  |  |  |  |  |
| Histology |  |  |  |  |  |  |  |
| Non-squamous | 301/684 | 44.0 (39.1–49.2) | 0.5997 | - | - | - | - |
| Squamous | 99/215 | 46.0 (37.4-56.1) |  |  |  |  |  |
| ECOG PS |  |  |  |  |  |  |  |
| ≥2 | 36/143 | 25.2 (17.6–34.8) | <0.0001 | 0.9580 | 0.2080 | <0.0001 | 2.60 (1.73-3.91) |
| 0-1 | 364/756 | 48.1 (43.3-53.3) |  |  |  |  |  |
| SNC metastases |  |  |  |  |  |  |  |
| Yes | 70/164 | 42.7 (33.2–53.9) | 0.6060 | - | - | - | - |
| No | 330/735 | 44.9 (40.1-50.1) |  |  |  |  |  |
| Bone metastases |  |  |  |  |  |  |  |
| Yes | 92/272 | 33.8 (27.2–41.4) | <0.0001 | 0.5626 | 0.1544 | 0.0003 | 1.75 (1.29-2.37) |
| No | 308/627 | 49.1 (43.8–54.9) |  |  |  |  |  |
| Liver metastases |  |  |  |  |  |  |  |
| Yes | 46/131 | 35.1 (25.7–46.8) | 0.0195 | 0.3110 | 0.2033 | 0.1260 | 1.36 (0.91-2.03) |
| No | 354/768 | 46.1 (41.4–51.1) |  |  |  |  |  |
| Baseline Steroids |  |  |  |  |  |  |  |
| Yes | 61/204 | 29.9 (22.8–38.4) | <0.0001 | - | - | - | - |
| No | 339/695 | 48.7 (43.7–54.2) |  |  |  |  |  |
| Previous RT |  |  |  |  |  |  |  |
| (No) | 321/710 | 45.2 (40.4–50.4) | 0.1120 | - | - | - | - |
| Non palliative intent | 23/42 | 54.8 (34.7–82.1) |  |  |  |  |  |
| Palliative intent | 56/147 | 38.1 (28.8–49.4) | 0.1939# |  |  |  |  |

**Table 4:** Univariate and multivariate analyses of PFS. *Available for 731 patients, not used in the multivariate analysis of the overall study population; ECOG-PS (≥ 2 *vs*. 0-1), CNS metastases (yes *vs*. no), bone metastases (yes *vs*. no) and liver metastases (yes *vs*. no) were used as adjusting factors for PD-L1 analysis.

|  | Progression Free Survival |  |
| --- | --- | --- |
|  | Univariate Analysis | Multivariate Analysis |
| VARIABLE<br>(Comparator) | HR (95% CI); *p* - value | HR (95% CI); *p* - value |
| PD-L1 expression*<br>< 90% vs ≥ 90% | 1.29 (1.01–1.66); *p*=0.0487 | 1.17 (0.91-1.510); *p*=0.2155 |
| Smoking status<br>(Never smoker)<br>Former smoker<br>Current smoker | 0.57 (0.44-0.73); *p*<0.0001<br>0.65 (0.50-0.85); *p*=0.0016 | 0.62 (0.48-0.79); *p*=0.0002<br>0.77 (0.59-1.01); *p*=0.0532 |
| Sex<br>Male vs Female | 0.97 (0.82–1.15); *p*=0.7326 | - |
| Age<br>Elderly vs Non-elderly | 1.01 (0.85–1.17); *p*=0.9983 | - |
| Histology<br>Non-sq. vs Squamous | 1.01 (0.83–1.20); *p*=0.9805 | - |
| ECOG PS<br>≥2 vs 0-1 | 2.65 (2.20–3.21); *p*<0.0001 | 2.48 (2.05–3.01); *p*<0.0001 |
| SNC metastases<br>Yes vs No | 1.32 (1.07–1.61); *p*=0.0076 | 1.22 (0.99–1.49); *p*=0.0529 |
| Bone metastases<br>Yes vs No | 1.75 (1.48–2.06); *p*<0.0001 | 1.46 (1.23–1.74); *p*<0.0001 |
| Liver metastases<br>Yes vs No | 1.97 (1.62–2.41); *p*<0.0001 | 1.69 (1.38–2.08); *p*<0.0001 |
| Baseline Steroids<br>Yes vs No | 2.05 (1.72-2.45); *p*<0.0001 | - |
| Previous RT<br>(No)<br>Non palliative intent<br>Palliative intent | 0.66 (0.42-1.05); *p*=0.0827<br>1.39 (1.14-1.71); *p*=0.0013 | - |

**Table 5:** Univariate and multivariate analyses of OS. *Available for 731 patients, not used in the multivariate analysis of the overall study population; ECOG-PS (≥ 2 vs. 0-1), CNS metastases (yes vs. no), bone metastases (yes vs. no) and liver metastases (yes vs. no) were used as adjusting factors for PD-L1 analysis.

|  | Overall Survival |  |
| --- | --- | --- |
|  | Univariate Analysis | Multivariate Analysis |
| VARIABLE<br>(Comparator) | HR (95% CI); *p* - value | HR (95% CI); *p* - value |
| PD-L1 expression*<br>< 90% vs ≥ 90% | 1.51 (1.10–2.07); *p*=0.0093 | 1.41 (1.02–1.92); *p*=0.0346 |
| Smoking status<br>(Never smoker)<br>Former smoker<br>Current smoker | 0.61 (0.46-0.82); *p*=0.0013<br>0.72 (0.53-0.98); *p*=0.0355 | 0.69 (0.51-0.92); *p*=0.0131<br>0.86 (0.63-1.17); *p*=0.3433 |
| Sex<br>Male vs Female | 1.08 (0.88–1.32); *p*=0.4408 | - |
| Age<br>Elderly vs Non-elderly | 1.04 (0.86–1.26); *p*=0.6923 | - |
| Histology<br>Non-sq. vs Squamous | 1.03 (0.82–1.29); *p*=0.8051 | - |
| ECOG PS<br>≥2 vs 0-1 | 3.18 (2.58–3.92); *p*<0.0001 | 3.01 (2.43–3.72); *p*<0.0001 |
| SNC metastases<br>Yes vs No | 1.28 (1.01–1.63); *p*=0.0472 | 1.16 (0.91–1.47); *p*=0.2316 |
| Bone metastases<br>Yes vs No | 1.82 (1.50–2.21); *p*<0.0001 | 1.53 (1.25–1.88); *p*<0.0001 |
| Liver metastases<br>Yes vs No | 1.96 (1.55–2.45); *p*<0.0001 | 1.66 (1.31–2.10); *p*<0.0001 |
| Baseline Steroids<br>Yes vs No | 2.43 (1.99-2.96); *p*<0.0001 | - |
| Previous RT<br>(No)<br>Non palliative intent<br>Palliative intent | 0.29 (0.13-0.67); *p*=0.0033<br>1.37 (1.07-1.72); *p*=0.0104 | - |

**Figure 2:**
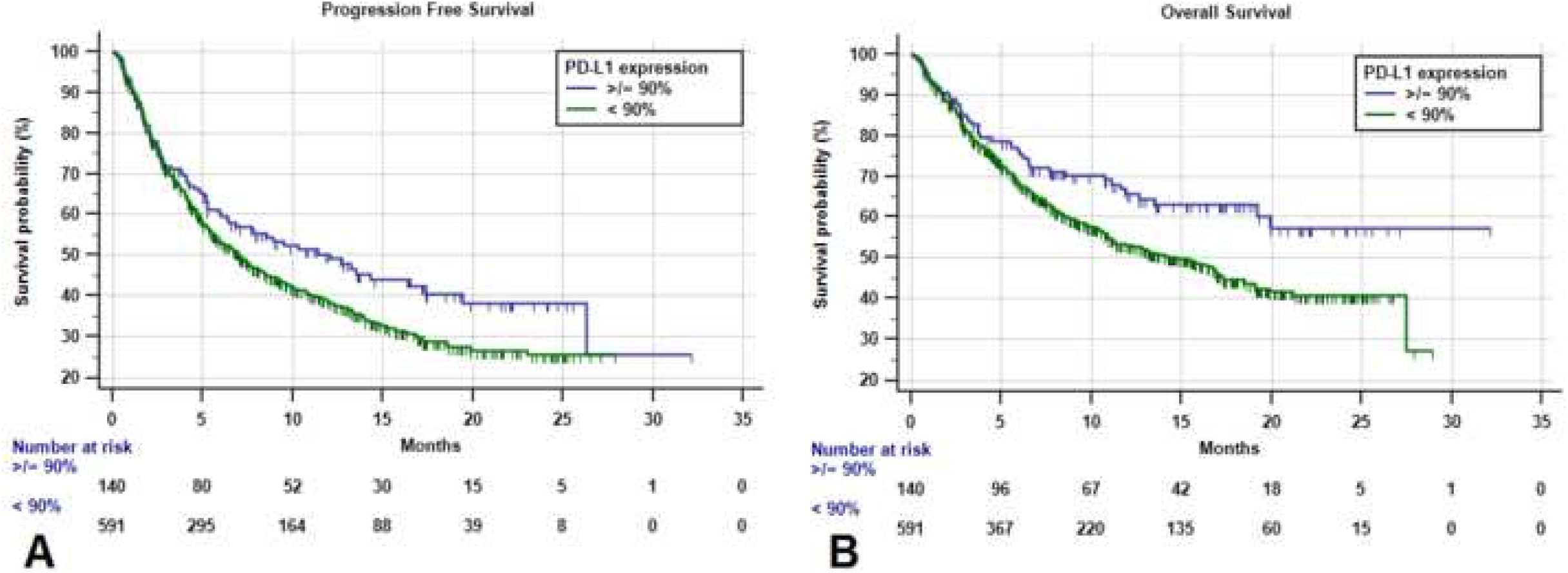
Kaplan-Meier survival curves of PFS (A) and OS (B) according to the computed PD-L1 TPS optimal cut offs.

At the univariate analysis baseline steroids and previous palliative RT were significantly related to a shortened PFS (Table 4). Only former smokers were confirmed to have a significantly prolonged PFS compared to never smoker patients at the multivariate analysis (Table 4). ECOG-PS, bone metastases and liver metastases were also confirmed to be independent predictors of shortened PFS (Table 4). At the univariate analysis baseline steroids and previous palliative RT were significantly related to a shortened OS. On the other hand, previous non-palliative RT was significantly related to a prolonged OS. At the multivariate analysis, former smokers were confirmed to have a significantly prolonged OS compared to never smoker patients, in contrast to what reported for current smokers. Even in this case, ECOG-PS, bone metastases and liver metastases were confirmed to be independent predictors of a shortened OS (Table 5). Figure 3 reported the Kaplan-Meier survival curves of PFS and OS for selected key subgroups. The median PFS of current smokers, former smokers and never smokers was 7.2 months (95%CI: 5.7-10.2; 205 events), 9.5 months (95%CI: 8.01-11.6; 316 events) and 4.1 months (95%CI: 3.3-5.7; 78 events), respectively (Figure 3A). Median OS of current smokers, former smokers and never smokers was 16.9 months (95%CI: 13.1-21.2; 199 censored patients), 19.9 months (95%CI: 16.8-27.5; 350 censored patients) and 9.4 months (95%CI: 6.9-15.0; 49 censored patients), respectively (Figure 3B). Median PFS of patients who received previous palliative RT, non-palliative RT and patients who did not received previous RT was 4.8 months (95%CI: 3.3-6.9; 115 events), 17.4 months (95%CI: 6.2-20.1; 19 events) and 8.4 months (95%CI: 7.3-10.2; 465 events), respectively (Figure 3C). Median OS of patients who received previous palliative RT, non-palliative RT and patients who did not received previous RT was 13.4 months (95%CI: 8.6-21.2; 82 censored patients), not reached (38 censored patients) and 17.2 months (95%CI: 15.2 – 19.9; 478 censored patients), respectively (Figure 3D). Supplementary Figure S1 reported the Kaplan-Meier survival curves of PFS and OS according to baseline CNS, bone and liver metastases.

**Figure 3:**
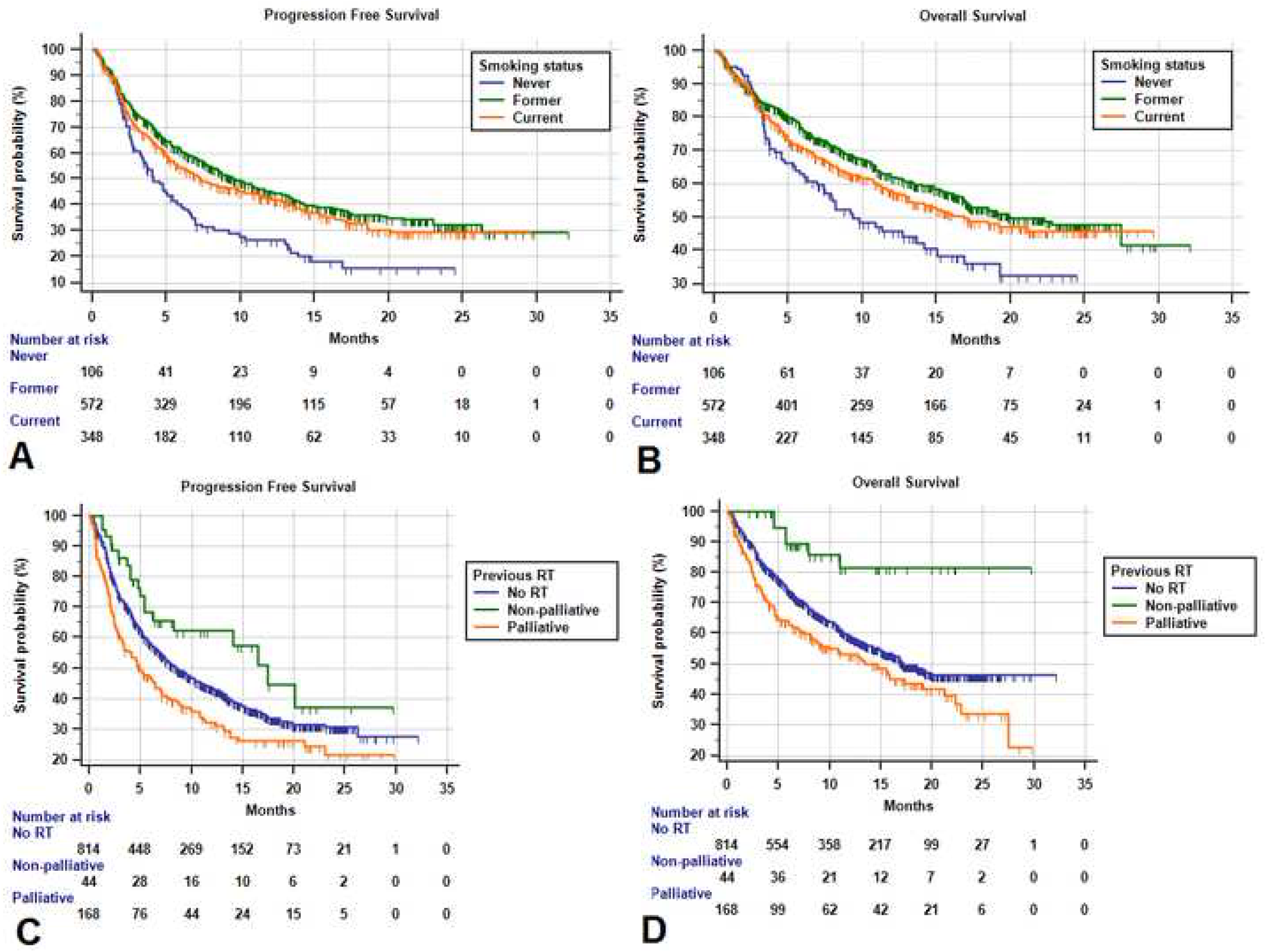
Kaplan-Meier survival curves according to the smoking status of PFS (A) and OS (B), and according to previous RT of PFS (C) and OS (D).

## Discussion

In this multicentre, real-life study we reported an ORR of 44.5%, a median PFS of 7.9 months and a median OS of 17.2 months (median follow-up of 14.6 months) to 1^st^ line pembrolizumab among patients with newly diagnosed advanced NSCLC and a PD-L1 expression of ≥50%. While the ORR in our study is similar to the ORR of 44.8% reported in the Keynote 024 study, the median PFS and OS observed in our population are shorter, which is compatible with the fair representation of patients with a PS of ≥ 2 and untreated brain metastasis, which is common in real life clinical practice [1-3]. The subgroup analysis of patients with PD-L1 TPS ≥ 50% receiving pembrolizumab (compared to standard platinum-based chemotherapy) within the Keynote-042 trial, otherwise reported an ORR of 39%, a PFS of 7.1 months and a median OS of 20.0 months (median follow-up of 12.8 months), which were slightly worse compared to our study population in terms of ORR [25]. Our efficacy results are also comparable to recent real-life studies [4-6]; however, to properly evaluate the comparability with clinical trials, we must consider some several key differences in the study population, beside the significant differences regarding the reported follow-up. Consistently with other recent retrospective analysis [4-6], also I our study patients with an ECOG-PS ≥ 2 (15.4%) were included, differently from the Keynote-024 trial [1]. Of note, patients with an ECOG-PS ≥ 2 are known to be not the best candidates for single agent immunotherapy [7]. ECOG-PS was indeed confirmed to be an independent predictor of worsened clinical outcome at each multivariate analysis in our study.

Recent network meta-analyses revealed that in NSCLC patients with PD-L1 TPS ≥ 50%, the addition of pembrolizumab to first line chemotherapy might have beneficial results in terms of ORR and PFS, compared to single agent pembrolizumab, apparently without any OS advantage [26-27]. However, these data were derived from clinical trials, therefore their reproducibility remains limited. Other differences in the study populations regard the exclusion from the Keynote-024 of oncogene-addicted patients, patients with untreated baseline CNS metastases, and patients requiring high dose steroids overall (both for cancer-related and unrelated indications). Data regarding baseline bone and liver metastases, which were already known to negatively affect immunotherapy clinical outcomes in NSCLC patients [28-29], were not provided within the Keynote-024 trial population. However, our results confirmed that both these metastatic sites were independently related to worse ORR (only bone metastases), PFS and OS, also in the first line setting of patients with PD-L1 TPS ≥ 50% treated with pembrolizumab monotherapy. The proportion of the deceased patients who did not received 2^nd^ line treatments (77.6%), and of the patients who discontinued pembrolizumab receiving a second line treatment (31.8%), reflects that the study population has been treated outside of clinical trials. Differently, 49.1% of the patients who discontinued pembrolizumab in the experimental arm of the Keynote-024 trial received a further disease-oriented treatment [2].

The ROC analysis for PD-L1 TPS of ORR of the Keynote-001 trial population, reported a good diagnostic ability within the range 1%-100%, in identifying NSCLC patients to be treated with single agent pembrolizumab, establishing 50% as PD-L1 TPS cut off (Youden’s J statistics between 45% and 50%) [30]. On the other hand, the ROC curve analysis for ORR did not identify a strong cut-off of PD-L1 expression within the range 50%-100%, to discriminate responders vs non-responders. Importantly, when used the recursive partition algorithm, we were able to identify the same cut-off of 90% recently reported by Aguilar et al [4], that discriminated patients who were more significantly more likely to have improved ORR, PFS and OS to pembrolizumab monotherapy, at univariate analysis. Of note, the multivariate analysis confirmed the 90% PD-L1 expression an independent predictor for improved ORR and OS.

The analysis of variance of PD-L1, revealed controversial results. It has already been reported that the smoking status could affect PD-L1 expression [31] and that liquid-based cytology are reliable for PD-L1 TPS estimation [23]; intriguingly, we found a significant trend toward an increasing PD-L1 expression estimation across smoking categories, and a significant difference regarding the estimated mean PD-L1 expression according to the tissue specimen. Despite the harmonization studies [32-33], our findings might be related to the different clinical practice (and immunohistochemical assays) of the several centers involved in this study.

The smoking status was already found to be related to clinical outcome of NSCLC patients receiving immunotherapy [34]. A recent study of patients with PD-L1 TPS ≥ 50%, treated with several immune checkpoint inhibitors across multiple lines, revealed a significantly improved ORR and a non-statistically significant trend towards an improved PFS/duration of response for heavy/light smokers compared to never smokers [11]. The authors identified a higher median tumor mutational burden (TMB) among heavy smokers as the potential mechanisms driving the difference in the clinical outcomes. Consistently, we confirmed the significant association between improved PFS and OS and smoking in the first-line setting of PD-L1 high NSCLC patients. Interestingly, only former smokers were confirmed to have significantly longer PFS and OS at the multivariate analysis. Moreover, the net values of ORR, PFS and OS of former smokers were numerically higher compared to current smokers, and the respective adjusted OR/HRs (for the comparison with never smokers), were concordantly lower. This might be related to the global/functional benefit of smoking cessation, which might has positively affected the clinical outcomes, without impairing the TMB-gain related to the smoking habit.

Our efficacy results according to previous RT raise some questions which still need to be addressed. Palliative-RT was significantly related to shortened PFS and OS, while non-palliative RT was significantly related to a prolonged OS. We also noticed that the HRs for palliative and non-palliative RT were opposite in both the PFS and OS analyses. Recent evidences suggested the positive role of adding stereotactic RT preceding pembrolizumab in NSCLC patients [35], and a recent study have shown a negative shift of the balance between favourable and unfavourable immune-modulating effects of RT according to its intent (palliative vs non-palliative) [13]. However, while interpreting these results, we must take into account the significant correlation between previous RT and both CNS and bone metastases. In particular, bone metastases resulted to be a key negative prognostic factor and patients receiving palliative RT had the highest incidence of bone metastases (68.5%), while patients who received non-palliative RT the lowest (6.8%). On the other hand, the association with non-palliative RT, might explain the absence of a significant prognostic role of CNS metastases. The significant association with the disease burden is also likely to explain the association between corticosteroid administration and impaired immunotherapy efficacy. Accordingly, a recent study has reported that baseline steroids administered for non-cancer related indication were not related to worse outcomes in NSCLC patients receiving PD-1 checkpoint inhibitors [36]. Despite the data lack availability regarding the steroids indication in our dataset, we can assume that in most cases they were administered for symptoms palliation. Among the limitations of the present study, we must cite the retrospective design, which exposes to selection biases, and the lack of centralized review (histological and imaging).

## Conclusion

In this study we confirmed the efficacy of first line single agent pembrolizumab in metastatic NSCLC patients with PD-L1 expression of ≥ 50% in a large real-life cohort, and confirmed the significant association of the smoking status and non-palliative RT, with improved clinical outcomes, establishing them as key features to be investigated in prospective clinical trials. Questions regarding the clinical efficacy in clinical subgroups, such as patients with poorer PS and with liver/bone metastases, still remains to be addressed. In particular, whether adding chemotherapy to pembrolizumab in these categories or not, in case of PD-L1 expression of ≥ 50%, remains to be determined.

## Data Availability

the datasets used during the present study are available from the corresponding author upon reasonable request.

## Acknowledgements

A special thanks to the “Consorzio Interuniversitario Nazionale per la Bio-Oncologia” for their support in this study

## Ethics approval and consent to participate

All patients provided written, informed consent to treatment with immunotherapy. The procedures followed were in accordance with the precepts of Good Clinical Practice and the declaration of Helsinki. The study was approved by the respective local ethical committees on human experimentation of each institution, after previous approval by the coordinating center (Comitato Etico per le provice di L’Aquila e Teramo, verbale N.15 del 28 Novembre 2019).

## Authors’ contributions

All authors contributed to the publication according to the ICMJE guidelines for the authorship (study conception and design, acquisition of data, analysis and interpretation of data, drafting of manuscript, critical revision). All authors read and approved the submitted version of the manuscript (and any substantially modified version that involves the author’s contribution to the study). Each author have agreed both to be personally accountable for the author’s own contributions and to ensure that questions related to the accuracy or integrity of any part of the work, even ones in which the author was not personally involved, are appropriately investigated, resolved, and the resolution documented in the literature.

## Funding

no funding was received.

## Consent for publication

Not applicable.

## Conflicts of Interest

Dr Alessio Cortellini received speaker fees and grant consultancies by Astrazeneca, MSD, BMS, Roche, Novartis, Istituto Gentili, Astellas and Ipsen. Dr Emilio Bria received speaker and travel fees from MSD, Astra-Zeneca, Pfizer, Helsinn, Eli-Lilly, BMS, Novartis and Roche. Dr Emilio Bria received grant consultancies by Roche and Pfizer. Dr. Marcello Tiseo received speaker fees and grant consultancies by Astrazeneca, Pfizer, Eli-Lilly, BMS, Novartis, Roche, MSD, Boehringer Ingelheim, Otsuka, Takeda and Pierre Fabre. Dr. Alessandro Morabito received speaker fees by Astra, Roche, BMS, MSD, Boehringer, Pfizer, Takeda. Dr Francesca Mazzoni received grant consultancies by MSD and Takeda. Dr Raffaele Gisti received speaker fees and grant consultancies by Astrazeneca and Roche. Dr Francesco Passiglia received grant consultancies by MSD and Astrazeneca. Dr Paolo Bironzo received grant consultancies by Astrazeneca and Boehringer-Ingelheim. Dr Alex Friedlaender received grant consultancies by Roche, Pfizer, Astellas and BMS. Dr Alfredo Addeo received grant consultancies by Takeda, MSD, BMJ, Astrazeneca, Roche and Pfizer. Dr Rita Chiari received speaker fees by BMS, MSD, Takeda, Pfizer, Roche and Astrazeneca. Dr Carlo Genova received speaker fees/grant consultancies by Astrazeneca, BMS, Boehringer-Ingelheim, Roche and MSD.

## Tables/Figures legend

**Supplementary Figure S1:** Kaplan-Meier survival curves according to baseline CNS, bone and liver metastases. Median PFS of patients with and without baseline CNS metastases was 5.9 months (95%CI: 3.9 – 7.1; 115 events) and 8.6 months (95%CI: 7.5 – 10.2; 484 events), respectively (S1A). Median OS of patients with and without baseline CNS metastases was 15.0 months (95%CI: 9.6 – 22.3; 99 censored patients) and 18.5 months (95%CI: 16.1 – 27.5; 499 censored patients), respectively (S1B). Median PFS of patients with and without baseline bone metastases was 4.5 months (95%CI: 3.4 – 5.7; 224 events) and 11.1 months (95%CI: 9.2 – 13.4; 375 events), respectively (S1C). Median OS of patients with and without baseline bone metastases was 10.9 months (95%CI: 8.1 – 12.8; 155 censored patients) and 27.5 months (95%CI: 18.5 – 27.5; 443 censored patients), respectively (S1D). Median PFS of patients with and without baseline liver metastases was 3.7 months (95%CI: 2.5 – 4.9; 125 events) and 9.8 months (95%CI: 8.0 – 11.3; 474 events), respectively (S1E). Median OS of patients with and without baseline liver metastases was 8.2 months (95%CI: 5.7 – 11.1; 62 censored patients) and 19.9 months (17.1 – 27.5; 536 censored patients), respectively (S1F).

**Supplementary file 1:** List of the oncological institution of the study

**Supplementary Table.** (for online use only)

| Institution | Department |
| --- | --- |
| St. Salvatore Hospital, University of L’Aquila, L’Aquila | Medical Oncology |
| SS Annunziata Hospital, Chieti | Medical Oncology |
| University Hospital of Parma, Parma | Medical Oncology |
| St. Camillo Forlanini Hospital, Rome | Pulmonary Oncology |
| University Hospital of Modena, Modena | Medical Oncology |
| S Maria Goretti Hospital, Latina | Medical Oncology |
| St. Andrea Hospital, Rome | Medical Oncology |
| Fondazione IRCCS Ca' Granda Ospedale Maggiore Policlinico, Milan | Medical Oncology |
| Campus Bio-Medico University, Rome | Medical Oncology |
| “Ospedali Riuniti” Hospital, Ancona | Medical Oncology |
| Policlinico Umberto I, Rome | Medical Oncology |
| Clinical Cancer Centre "Giovanni Paolo II", Bari | Thoracic Oncology Unit |
| Hospital of Fabriano, Fabriano | Medical Oncology |
| “Augusto Murri” Hospital, Fermo | Medical Oncology |
| St. Gerardo Hospital, Monza | Medical Oncology |
| IRCCS – Istituto Nazionale Tumori, Fondazione “G. Pascale”, Napoli | Medical Oncology |
| IRCCS Sacro Cuore Don Calabria, Negrar | Medical Oncology |
| University Hospital of Udine, Udine | Medical Oncology |
| ASST-Sette Laghi, Varese | Medical Oncology |
| University Hospital “A.Gemelli”, Rome | Comprehensive Cancer Center |
| “Madre Teresa Di Calcutta” Hospital Padova Sud, Monselice | Medical Oncology |
| Hospital of Macerata | Medical Oncology |
| “F. Spaziani” Hospital, Frosinone | Medical Oncology |
| “Careggi” University Hospital, Florence | Medical Oncology |
| AUSL Romagna, Ravenna | Department of Oncology and Hematology |
| “Monaldi” Hospital, Naples | Pneumo-Oncology Unit |
| Erasmus Medical Center, Rotterdam, the Netherlands | Department of Pulmonary Diseases |
| “San Luigi-Gonzaga” University Hospital, Orbassano | Department of Oncology |
| “Fondazione IRCCS Istituto Nazionale dei Tumori”, Milan | Department of Medical Oncology |
| “Santa Maria della Misericordia” Hospital, Perugia | Medical Oncology |
| University Hospital of Geneva, Geneva | Medical Oncology |
| United Lincolnshire Hospital Trust, Lincoln | Medical Oncology |

## References

1. Reck M, Rodríguez-Abreu D, Robinson AG, et al. Pembrolizumab versus Chemotherapy for PD-L1-Positive Non-Small-Cell Lung Cancer. N Engl J Med. 2016 Nov 10;375(19):1823–1833. Epub 2016 Oct 8.

2. Reck M, Rodríguez-Abreu D, Robinson AG, et al. Updated Analysis of KEYNOTE-024: Pembrolizumab Versus Platinum-Based Chemotherapy for Advanced Non-Small-Cell Lung Cancer With PD-L1 Tumor Proportion Score of 50% or Greater. J Clin Oncol. 2019 Mar 1;37(7):537–546. doi: 10.1200/JCO.18.00149.

3. Reck M, Rodríguez-Abreu D, Robinson AG, et al. OA14.01 KEYNOTE-024 3-Year Survival Update: Pembrolizumab vs Platinum-Based Chemotherapy for Advanced Non– Small-Cell Lung Cancer. J Thorac Oncol 2019. Volume 14, Issue 10, S243

4. Aguilar EJ, Ricciuti B, Gainor JF, et al. Outcomes to first-line pembrolizumab in patients with non-small-cell lung cancer and very high PD-L1 expression. Ann Oncol. 2019 Oct 1;30(10):1653–1659. doi: 10.1093/annonc/mdz288.

5. Tamiya M, Tamiya A, Hosoya K, et al. Efficacy and safety of pembrolizumab as firstline therapy in advanced non-small cell lung cancer with at least 50% PD-L1 positivity: a multicenter retrospective cohort study (HOPE-001). Invest New Drugs. 2019 Dec;37(6):1266–1273. doi: 10.1007/s10637-019-00843-y. Epub 2019 Aug 7.

6. Velcheti V, Chandwani S, Chen X, et al. Outcomes of first-line pembrolizumab monotherapy for PD-L1-positive (TPS ≥50%) metastatic NSCLC at US oncology practices. Immunotherapy. 2019 Dec;11(18):1541–1554. doi: 10.2217/imt-2019-0177. Epub 2019 Nov 27.

7. Facchinetti F, Mazzaschi G, Barbieri F, et al. First-line pembrolizumab in advanced non-small cell lung cancer patients with poor performance status. European J of Cancer 2020. In Press.

8. Gandhi L, Rodríguez-Abreu D, Gadgeel S, et al. Pembrolizumab plus Chemotherapy in Metastatic Non-Small-Cell Lung Cancer. N Engl J Med. 2018 May 31;378(22):2078–2092. doi: 10.1056/NEJMoa1801005. Epub 2018 Apr 16.

9. Paz-Ares L, Luft A, Vicente D, et al. Pembrolizumab plus Chemotherapy for Squamous Non-Small-Cell Lung Cancer. N Engl J Med. 2018 Nov 22;379(21):2040–2051. doi: 10.1056/NEJMoa1810865. Epub 2018 Sep 25.

10. Eisenhauer EA, Therasse P, Bogaerts J et al. New response evaluation criteria in solid tumours: revised RECIST guideline (version 1.1). Eur J Cancer 2009; 45: 228–247.

11. Gainor JF, Rizvi H, Jimenez Aguilar E, et al. Clinical Activity of Programmed Cell Death 1 (PD-1) Blockade in Never, Light, and Heavy Smokers with Non-Small Cell Lung Cancer and PD-L1 Expresion ≥ 50%. Ann Oncol in press. https://doi.org/10.1016/j.annonc.2019.11.015

12. Gridelli C, Balducci L, Ciardiello F, et al. Treatment of Elderly Patients With Non-Small-Cell Lung Cancer: Results of an International Expert Panel Meeting of the Italian Association of Thoracic Oncology. Clin Lung Cancer. 2015 Sep;16(5):325–33.

13. Bersanelli M, Lattanzi E, D’Abbiero N, et al. Palliative radiotherapy in advanced cancer patients treated with immune-checkpoint inhibitors: The PRACTICE study. Biomed Rep. 2020 Feb;12(2):59–67. doi: 10.3892/br.2019.1265.

14. Koletsi D, Pandis N. The chi-square test for trend. Am J Orthod Dentofacial Orthop. 2016 Dec;150(6):1066–1067. doi: 10.1016/j.ajodo.2016.10.001.

15. Bonate, P. L. (2017) Effect of correlation on covariate selection in linear and nonlinear mixed effect models. Pharmaceut. Statist., 16: 45–54. doi: 10.1002/pst.1776.

16. >Hosmer DW Jr, Lemeshow S, Sturdivant RX Applied Logistic Regression. Third Edition. New Jersey: John Wiley & Sons (2013).

17. Kaplan EL, Meier P. Nonparametric estimation of incomplete observations. J. Am. Stat. Assoc. 1958. 53:457–481.

18. Schemper M, Smith TL. A note on quantifying follow-up in studies of failure time. Controlled Clinical Trials 1997. 17:343-346.10.

19. Cox DR. Regression models and life tables (with discussion). Journal of the Royal Statistical Society (Series B) 1972. 74: 187–200.

20. PD-L1 IHC 22C3 pharmDx Interpretation Manual – NSCLC. Available at https://www.agilent.com/cs/library/usermanuals/public/29158_pd-l1-ihc-22C3-pharmdx-nsclc-interpretation-manual.pdf. Last access on February 18th, 2020.

21. Hanley JA, McNeil BJ. The meaning and use of the area under a receiver operating characteristic (ROC) curve. Radiology. 1982 Apr;143(1):29–36.

22. Ihaka, R. and Gentleman, R. A language for data analysis and graphics. J Comput Graph Stat. 1996; 5: 299–314

23. Teixidó C, Vilariño N, Reyes R, et al. PD-L1 expression testing in non-small cell lung cancer. Ther Adv Med Oncol. 2018; 10: 1758835918763493. Published online 2018 Apr 11. doi: 10.1177/1758835918763493.

24. Howell, David (2002). Statistical Methods for Psychology. Duxbury. pp. 324–325. ISBN 0-534-37770-X.

25. Mok TSK, Wu YL, Kudaba I, et al. Pembrolizumab versus chemotherapy for previously untreated, PD-L1-expressing, locally advanced or metastatic non-small-cell lung cancer (KEYNOTE-042): a randomised, open-label, controlled, phase 3 trial. Lancet. 2019 May 4;393(10183):1819–1830. doi: 10.1016/S0140-6736(18)32409-7.

26. Zhou Y, Lin Z, Zhang X, et al. First-line treatment for patients with advanced non-small cell lung carcinoma and high PD-L1 expression: pembrolizumab or pembrolizumab plus chemotherapy. J Immunother Cancer. 2019 May 3;7(1):120. doi: 10.1186/s40425-019-0600-6.

27. Kim R, Keam B, Hahn S, et al. First-line Pembrolizumab Versus Pembrolizumab Plus Chemotherapy Versus Chemotherapy Alone in Non-small-cell Lung Cancer: A Systematic Review and Network Meta-analysis. Clin Lung Cancer. 2019 Sep;20(5):331-338.e4. doi: 10.1016/j.cllc.2019.05.009.

28. Landi L, D’Incà F, Gelibter A, et al. Bone metastases and immunotherapy in patients with advanced non-small-cell lung cancer. J Immunother Cancer. 2019 Nov 21;7(1):316. doi: 10.1186/s40425-019-0793-8.

29. Botticelli A, Salati M, Di Pietro FR, et al. A nomogram to predict survival in non-small cell lung cancer patients treated with nivolumab. J Transl Med. 2019 Mar 27;17(1):99. doi: 10.1186/s12967-019-1847-x.

30. Garon EB, Rizvi NA, Hui R, et al. Pembrolizumab for the treatment of non-small-cell lung cancer. N Engl J Med. 2015 May 21;372(21):2018–28. doi: 10.1056/NEJMoa1501824.

31. Koh J, Go H, Keam B, et al. Clinicopathologic analysis of programmed cell death-1 and programmed cell death-ligand 1 and 2 expressions in pulmonary adenocarcinoma: comparison with histology and driver oncogenic alteration status. Mod Pathol 2015;28:1154-66. 10.1038/modpathol.2015.63

32. Adam J, Le Stang N, Rouquette I, et al. Multicenter harmonization study for PD-L1 IHC testing in non-small-cell lung cancer. Ann Oncol. 2018 Apr 1;29(4):953–958. doi: 10.1093/annonc/mdy014.

33. Marchetti A, Barberis M, Franco R, et al. Multicenter Comparison of 22C3 PharmDx (Agilent) and SP263 (Ventana) Assays to Test PD-L1 Expression for NSCLC Patients to Be Treated with Immune Checkpoint Inhibitors. J Thorac Oncol. 2017 Nov;12(11):1654–1663. doi: 10.1016/j.jtho.2017.07.031.

34. Kim JH, Kim HS, Kim BJ, et al. Prognostic value of smoking status in non-small-cell lung cancer patients treated with immune checkpoint inhibitors: a meta-analysis. Oncotarget. 2017 Jun 28;8(54):93149–93155. doi: 10.18632/oncotarget.18703.

35. >Theelen WSME, Peulen HMU, Lalezari F, et al. Effect of Pembrolizumab After Stereotactic Body Radiotherapy vs Pembrolizumab Alone on Tumor Response in Patients With Advanced Non-Small Cell Lung Cancer: Results of the PEMBRO-RT Phase 2 Randomized Clinical Trial. JAMA Oncol. 2019 Jul 11. doi: 10.1001/jamaoncol.2019.1478.

36. Ricciuti B, Dahlberg SE, Adeni A, et al. Immune Checkpoint Inhibitor Outcomes for Patients With Non-Small-Cell Lung Cancer Receiving Baseline Corticosteroids for Palliative Versus Nonpalliative Indications. J Clin Oncol. 2019 Aug 1;37(22):1927–1934. doi: 10.1200/JCO.19.00189.

